# Clinical genetic risk variants inform a functional protein interaction network for tetralogy of Fallot

**DOI:** 10.1101/2021.02.17.21251707

**Authors:** Miriam S. Reuter, Rajiv R. Chaturvedi, Rebekah K. Jobling, Giovanna Pellecchia, Omar Hamdan, Wilson W.L. Sung, Thomas Nalpathamkalam, Pratyusha Attaluri, Candice K. Silversides, Rachel M. Wald, Christian R. Marshall, Simon Williams, Bernard D. Keavney, Bhooma Thiruvahindrapuram, Stephen W. Scherer, Anne S. Bassett

## Abstract

**Background:** Tetralogy of Fallot (TOF), the most common cyanotic heart defect in newborns, has evidence of multiple genetic contributing factors. Identifying variants that are clinically relevant is essential to understand patient-specific disease susceptibility and outcomes, and could contribute to delineating pathomechanisms.

**Methods and Results:** We used a clinically-driven strategy and current guidelines to re-analyze exome sequencing data from 811 probands with TOF, focused on identifying rare loss-of-function and other likely pathogenic variants in congenital heart disease (CHD) genes. In addition to confirming a major contribution of likely pathogenic variants in *FLT4* (VEGFR3; n=14) and *NOTCH1* (n=11), we identified 1-3 such variants in each of 21 other CHD genes, including *ATRX, DLL4, EP300, GATA6, JAG1, NF1, PIK3CA, RAF1, RASA1, SMAD2*, and *TBX1*. There were also three emerging CHD/TOF candidate genes with multiple loss-of-function variants in this cohort: *KDR* (n=4), *IQGAP1* (n=3), and *GDF1* (n=8). In total, these variants were identified in 64 probands (7.9%). Using the 26 composite genes in a STRING protein interaction enrichment analysis revealed a biologically relevant network (p-value 3.3e-16), with VEGFR2 (*KDR*) and NOTCH1 representing central nodes. Variants associated with arrhythmias/sudden death and/or heart failure indicated factors that could influence long-term outcomes.

**Conclusions:** The results are relevant to precision medicine for TOF. They suggest considerable clinical yield from genome-wide sequencing, and further evidence for *KDR* as a CHD/TOF gene and VEGF and Notch signaling as mechanisms in human disease. Harnessing genetic heterogeneity of single gene defects could inform etiopathogenesis and help prioritize novel candidate genes for TOF.

## Introduction

Tetralogy of Fallot (TOF) affects about one in 3000 live births, and is the most common cyanotic heart defect in newborns ^1, 2^. Initially described in 1671 by Danish anatomist Niels Stensen, the four components (pulmonary outflow tract obstruction, aorta overriding both ventricles, ventricular septal defect, and hypertrophy of the right ventricle) represent a single developmental anomaly ^3, 4^. Further detailed anatomical documentation by Arthur Fallot, Maude Abbott, and others, laid the foundation first for palliative, and later corrective, surgical procedures ^4, 5^. Advances in imaging, medical management, and surgeries across the lifespan have transformed TOF from a usually fatal pediatric condition to a chronic disease that is more prevalent in adults than children, with life expectancy into the seventh decade and beyond ^6-9^.

The pursuit of determining the genetic underpinnings and recognizing how these may affect late outcomes in TOF, has proceeded in parallel with these clinical advances ^10-13^. This research began with the recognition of multi-system syndromes in approximately 20% of patients, most commonly caused by 22q11.2 microdeletions, other copy number variants, or aneuploidies ^14^, along with some single-gene defects ^15^. Availability of newer genomic technologies, particularly genome-wide sequencing, has expanded gene discovery studies. The cumulative genetic evidence indicates a pattern of molecular etiology for TOF that is characterized by genetic heterogeneity and some distinction from congenital heart disease (CHD) as a whole ^14, 16-21^. However, there has been relatively limited consideration of the clinical pathogenicity of genetic variants for TOF and translation of findings into the clinic ^22, 23^.

The cohort studied here is the largest genome-wide sequencing dataset for TOF ^20, 24^. Previous research identified clusters of deleterious variants in *NOTCH1* and *FLT4* which surpassed statistical thresholds for genome-wide significance and were thus considered predisposing to TOF in almost 7% of the probands; a potential association with *TBX1* deleterious variants was also reported ^20^. However, many of the *NOTCH1* and *FLT4* variants constituted missense variants of uncertain significance, according to standard American College of Medical Genetics (ACMG) adjudication guidelines (details on *TBX1* variants were not provided) ^25^. Besides including abundant variants of uncertain significance, genome-wide variant burden analyses often lack power to detect rare gene-disease associations, which contribute largely to the genetic heterogeneity, and form the basis for defining clinically reportable genetic variants.

The objective of this study was to improve our understanding of variants that are of relevance to clinical practice for patients with TOF. We used ACMG interpretation guidelines to re-analyze the exome sequencing data of 811 probands with TOF, and focused on delineating variants that were considered relevant for the congenital cardiac phenotype. We identified 64 (7.9%) of 811 pediatric TOF probands to have pathogenic/likely pathogenic variants in known CHD genes (n = 50), or loss-of-function variants in emerging CHD/TOF candidate genes (n = 15; Figure 1). The implicated genes encode proteins that functionally interact, indicating that the heterogeneous genetic architecture could inform mechanism. Other pathogenic/likely pathogenic variants identified add to potential genetic implications for cardiovascular outcomes.

**Figure 1:**
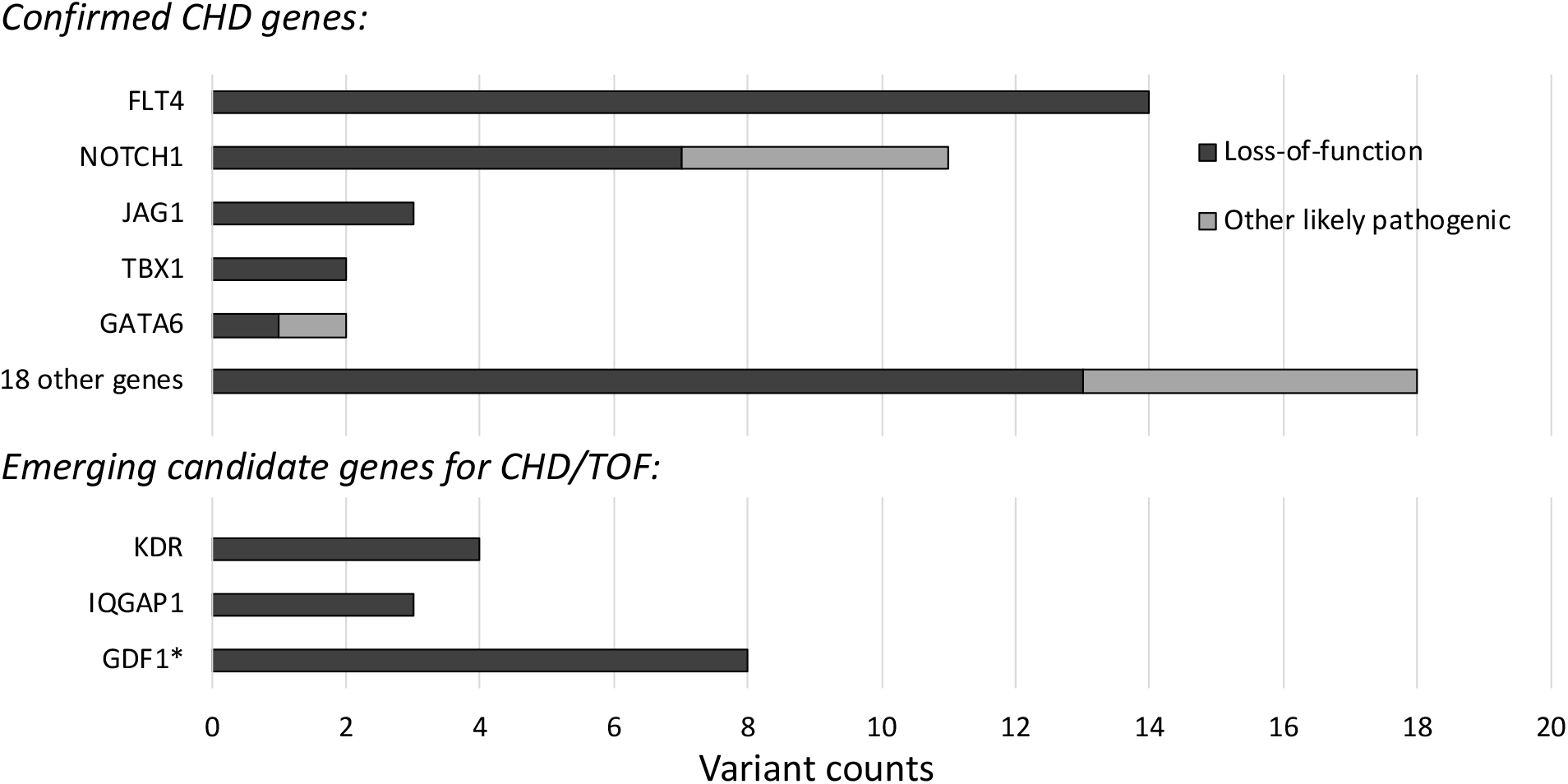
Clinically relevant variants associated with congenital heart disease (CHD), identified in exome sequencing data of n = 811 probands with tetralogy of Fallot (TOF). Likely pathogenic loss-of-function and missense variants meeting ACMG criteria for clinical relevance (pathogenic or likely pathogenic, n = 50) in 23 recognized CHD genes were identified in 49 probands. Loss-of-function variants (n = 15) in 3 emerging candidate genes for CHD/TOF were identified in another 15 probands. Variant and gene details are provided in Tables S2 and S3, respectively, and Figure S1 provides details of *KDR* variants. Additional variants of uncertain significance are provided in Table S5. * Note: Comprehensive assessment of gene *GDF1* was not possible due to insufficient coverage of this locus with the exome sequencing data available.

## Methods

We studied the exome sequencing data of a multi-centre cohort of probands with TOF, accessed through the European Genome-phenome Archive (EGA ^26^; https://www.ebi.ac.uk/ega), study accession EGAS00001003302 ^20^. The cohort was reported to be of Northern European ancestry, not to exhibit features of recognized malformation or developmental syndromes ^27^, and to have previously excluded 22q11.2 microdeletions by multiplex ligation-dependent probe amplification ^20^. We downloaded 829 bam files from EGA, subsequently excluding 18 files that were corrupted, aligned to a different (bwa VN:0.7.6a-r433, b37) reference genome, or duplicates (supplementary information).

For the analyses presented here, we considered n = 811 unique exome sequencing datasets (Table S1), aligned to GRCh37 (hg19). The data were analyzed at The Centre for Applied Genomics (TCAG, The Hospital for Sick Children, Toronto, Canada) under a research protocol of the Hospital for Sick Children (REB# 0019980189). We prioritized the dataset for rare variants (substitutions and small insertions/deletions) affecting genes associated with congenital heart defects (https://omim.org/, http://chdgene.victorchang.edu.au/, https://pubmed.ncbi.nlm.nih.gov/; as of September 2020), and other cardiac conditions putatively relevant to outcome (supplementary information). For the yield of pathogenic/likely pathogenic variants, we pursued a conservative interpretation strategy as per ACMG consensus guidelines ^25^. Variants covered by less than 10x were omitted, and all reported variants were visualized using IGV (http://software.broadinstitute.org/software/igv/). For candidate genes *KDR* and *IQGAP1*, we compared the loss-of-function variant counts to those identified by genome sequencing of two control cohorts: (i) participants in the 1000 genomes project (n = 2,504 genomes, all ancestries; https://www.internationalgenome.org/), and (ii) parents of European ancestry from the Autism Speaks MSSNG database (n = 3,697 genomes in DB6; https://www.mss.ng/). The prevalence of the *GDF1* stopgain variant p.Cys227* was only compared to the n = 3,697 European controls from MSSNG, as this variant is known to be more common among Europeans. Functional protein interaction analyses were performed with Cytoscape ^28^ software v.3.8.2, using the STRING app ^29^.

## Results

Re-analyzing the exome sequencing data from 811 probands with TOF, we identified five CHD-associated genes with multiple pathogenic/likely pathogenic variants (Figure 1). These included 14 likely pathogenic loss-of-function variants in *FLT4* (all loss-of-function), and 11 in *NOTCH1* (7 loss-of-function, 4 missense) (Table S2). That we observed the highest prevalence for variants in these two genes, collectively 25/811 (3.1%), but lower than previously reported variant yields, was expected given findings from studies of this and other cohorts using different adjudication methods ^18, 20, 21^. We also identified three likely pathogenic variants in *JAG1* (OMIM-P 187500, 118450), and two each in *TBX1* (OMIM-P 187500), and *GATA6* (OMIM-P 187500, 600001) (Table S2). Consistent with the genetic heterogeneity of TOF, we identified 16 other individuals to have one pathogenic/likely pathogenic variant (11 loss-of-function, 5 missense) in 16 CHD genes: *ARHGAP31, ATRX, CACNA1C, CHD7, CSNK2A1, DLL4, EP300, GATAD2B, KAT6A, LZTR1, NF1, NODAL, PIK3CA, RAF1, RASA1*, and *SMAD2*, and one individual with loss-of-function variants in two genes, *ASXL1* and *PSMD12* (Table S2). Thus, there were 49 probands in this cohort identified to have at least one pathogenic/likely pathogenic variant in a CHD-associated gene (49/811; 6.0%).

In three emerging CHD candidate genes (*KDR, IQGAP1*, and *GDF1*), i.e., with substantial research evidence but as yet insufficient to clinically deem variants “likely pathogenic”, we identified multiple loss-of-function variants in 16 individuals in this cohort (Figure 1, Table S3). Four individuals with TOF had high-confidence loss-of-function variants in *KDR*, compared with none in 6,201 controls (Fisher’s exact test (FET) *p* = 1.8E-4; gnomAD observed/expected loss-of-function constraint score (o/e LOF) = 0.15). The significant enrichment found for this large cohort adds further evidence for *KDR* as a TOF/CHD gene to that provided by several recent studies reporting rare loss-of-function variants in *KDR* in independent cohorts with TOF or other conotruncal defects ^18, 21, 30^ (Figure S1).

*IQGAP1* loss-of-function variants have also been reported in several cohorts of TOF and other CHD ^18, 31, 32^. In the TOF cohort analyzed here, there were three individuals with loss-of-function variants in *IQGAP1* with significant enrichment compared to 1000 genomes control data (3/811 vs. 1/2,504; FET: *p* = 4.8E-2; gnomAD o/e LOF = 0.19), but at the non-significant trend level compared to control data comprising parents in an autism cohort (3/811 vs. 5/3,697; FET: *p* = 1.6E-1). For *GDF1*, the pathogenicity of biallelic (homozygous or compound heterozygous) variants is well established [OMIM-P 208530] ^21^, and heterozygous variants have also been suggested as risk factors for CHD ^33^ [OMIM-P 613854]. In the current exome data, the prevalence of *GDF1* heterozygous loss-of-function variants is likely to be underestimated due to insufficient coverage of this locus. Nonetheless, we identified 8 individuals to have such a variant, including an identical, heterozygous *GDF1* stopgain variant p.Cys227* in 7 of the 706 unrelated probands having samples with adequate read depths (≥10x). In comparison, the carrier frequency of this variant was 6 of 3,697 European controls (FET: p = 1.9E-3), indicating significant enrichment of heterozygous loss-of-function variants in *GDF1* in the TOF cohort studied.

Taken together, we identified 64 probands (7.9%) with n = 50 pathogenic/likely pathogenic variants in 23 known CHD genes (Table S2) or n = 15 loss-of-function variants in 3 emerging CHD/TOF candidate genes (Table S3) in these TOF exome data. We note that the majority of these variants (41/65, 63.1%) and genes (23/26, 88.5%) were not previously reported for this cohort ^20^ (Tables S2, S3).

We next considered the 23 identified TOF disease genes that had clinically relevant variants and 3 emerging candidate genes as a group, in an *in silico* functional interaction analysis. This showed evidence for a highly interactive network of the encoded proteins (STRING interaction enrichment *p* value 3.3E-16). VEGFR2 (gene *KDR*) and NOTCH1 were central nodes within this network map, each connecting directly with 11 other proteins. In all, 23 of the 26 genes (88.5%) form an interactive network, related to VEGFR2 (*KDR*) or NOTCH1 by ≤2 edges (Figure 2). Pathogenic/likely pathogenic loss-of-function variants from an independent cohort of 424 children with TOF ^21^ supported, and slightly extended, this network, identifying NOTCH2 as an additional protein with multiple (five) interactions (Table S4, Figure S2). All of the composite genes are expressed in human hearts (https://www.proteinatlas.org/), but at varying levels (Figures 2, S2). A pathway enrichment map for those proteins is provided as supplemental material (Figure S3).

**Figure 2:**
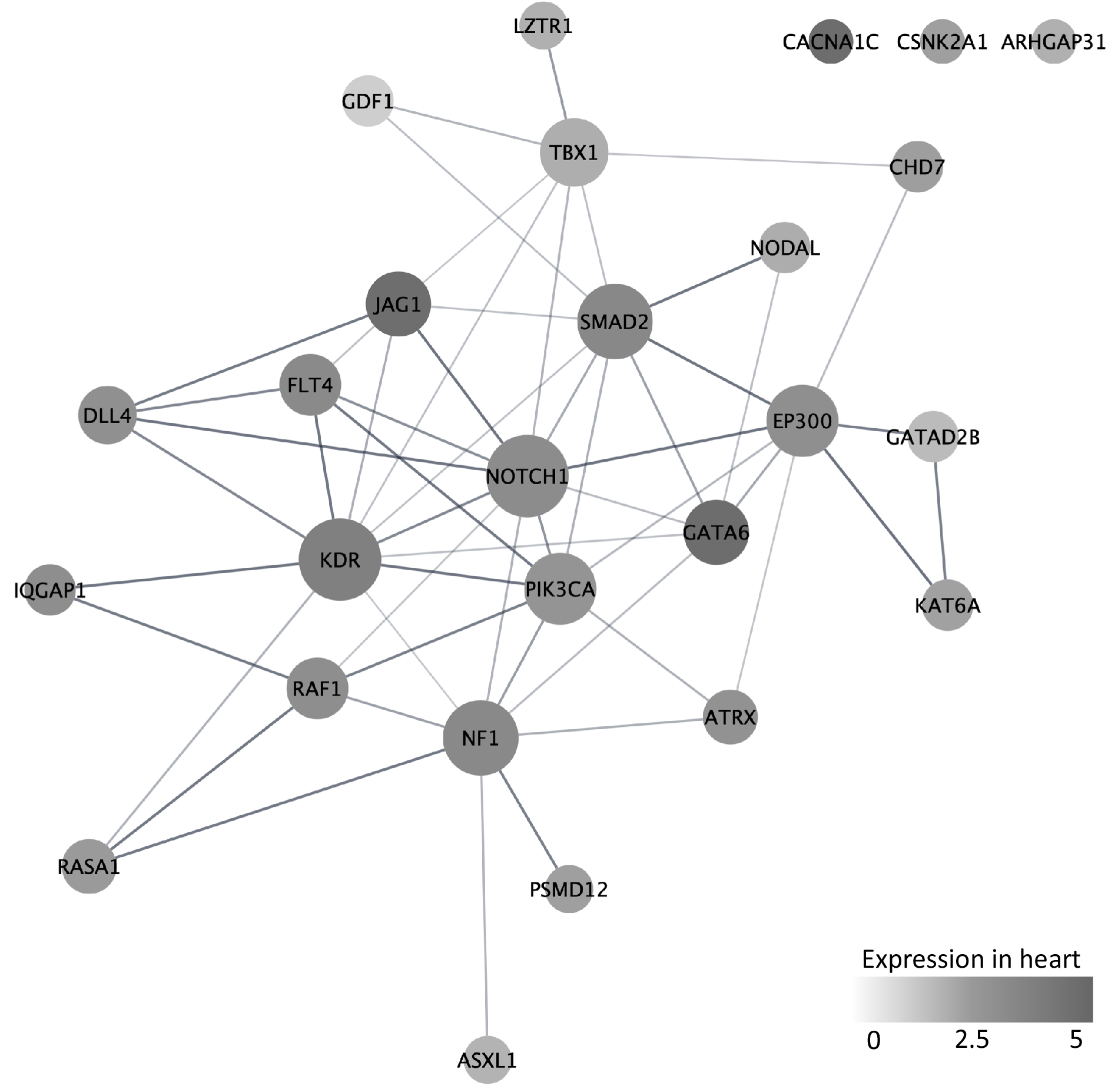
Functionally interacting proteins encoded by confirmed genes (n = 23) and emerging candidate genes (n = 3) for CHD/TOF identified in n = 811 individuals with TOF. Network analysis was performed using Cytoscape, STRING and the 26 genes identified from exome sequencing of this cohort (STRING interaction enrichment *p* value 3.3E-16; see text for details). Node sizes (circles) represent the connectivity (numbers of edges to other proteins). Node greyscales represent the degree of expression from the Gene Expression Omnibus (GEO) database (heart tissue). Edge widths represent the confidence (strength of data support). 23 of the 26 genes form an interactive network with VEGFR2 (gene *KDR*) and NOTCH1 as central nodes, each connecting directly with 11 other proteins. Figure S2 shows a slightly extended network (e.g., NOTCH2 with 5 interactions) after adding data from an independent TOF cohort. Not shown are four proteins of uncertain relevance to TOF, but with likely pathogenic variants observed in the sample (Table S6) that connect to the network: GLI2 (NOTCH1, SMAD2, JAG1, PSMD12), APC (NOTCH1, IQGAP1, PSMD12, CSNK2A1), TCF12 (NOTCH1, SMAD2, EP300), POLR1A (EP300).

Notably, we additionally identified rare nonsynonymous or predicted splice-altering variants of uncertain significance, according to ACMG interpretation guidelines. In some cases, clinical data or knowledge on the parental genotypes, which were inaccessible for this study, could inform more accurate variant classifications. The majority of *NOTCH1* rare nonsynonymous missense and in-frame variants that were previously reported for this sample ^20^ were variants of uncertain significance (n = 21; Table S5). Three of these variants – p.(Glu1294Lys), p.(Gly200Arg) ^34^, and p.(Pro143Leu) – were identified in probands with other variants identified as likely pathogenic: *NOTCH1* p.(Gln1733*), *NF1* c.5206-1G>C, and *EP300* p.(Phe1595Val), respectively (Table S5), representing examples of within-individual allelic or genetic heterogeneity. There were other CHD-relevant genes or candidates with variants of uncertain significance, including loss-of-function variants in *CHD4, ECE1, SMAD6, ZFPM1, PRKD1* and *VEGFA* (Table S5).

We also report pathogenic/likely pathogenic variants for childhood-onset disorders, but with less established evidence at this time for clinical relevance to TOF/CHD (Table S6). These include loss-of-function variants in *POLR1A* (2x), *TCF12* (2x), *APC*, and *GLI2* (Table S6) where, interestingly, the encoded proteins link to the network map for TOF (Figure 2).

To further investigate the potential clinical utility of exome sequencing, we also interrogated the dataset for rare variants with potential clinical implications for cardiovascular management and outcome. In 16 (2.0%) of the 811 probands we identified pathogenic/likely pathogenic variants meeting these criteria. There were 11 variants associated with cardiac hypertrophy, arrhythmia and sudden cardiac death: hypertrophic cardiomyopathy (*MYBPC3* (3x), *MYH7, MYL2, TNNI3*), arrhythmogenic right ventricular dysplasia (*DSP* (2x), *DSC2*), Brugada syndrome (*SCN5A*), and dilated cardiomyopathy (*DMD*) (Table S7). There were also five variants in genes (*LZTR1, RAF1, CACNA1C, NF1, RASA1*) implicated in other conditions, including e.g., Noonan and Timothy syndromes ^35, 36^, that in addition have been reported to be associated with CHD, thus were considered in the etiologic analysis (Tables S2, S7).

## Discussion

Precision medicine is an emerging concept that involves health management based on individual characteristics, including genetic disease susceptibilities and pharmacogenomics ^37, 38^. Delineation of disease-causing and very high impact variants and genes and their functional networks will advance both precision health initiatives and our understanding of the relevant molecular mechanisms. The rationale for re-analyzing this cohort of 811 individuals with TOF was to focus on identifying sequence variants with sufficient evidence for pathogenicity, according to consensus clinical guidelines ^25^, that could inform disease etiologies of TOF. We also aimed to detect rare and emerging gene-disease associations, that had not been noted in previous genome-wide enrichment studies of the cohort ^20^. We identified 50 pathogenic/likely pathogenic variants for TOF, plus 15 loss-of-function variants in emerging CHD/TOF candidate genes. The majority of variants (41/65, 63.1%) and genes (23/26, 88.5%) were not previously reported for this cohort (Tables S2, S3). Collectively, the results document both genetic and allelic heterogeneity at the clinical level in the pathogenesis of TOF across this large cohort, and in some cases within individual. The findings may also help to define minimal clinical gene panels for TOF.

Likely pathogenic loss-of-function variants in *FLT4* (VEGFR3), *NOTCH1*, and *JAG1* were among the most common findings, further confirming a major role for VEGF and Notch signaling in TOF. A dosage effect of VEGF/Notch signaling in outflow tract development and TOF has long been suggested from mouse models ^39^, and has been recently substantiated by several human genetic studies ^18, 20, 32^. We also provide further evidence that haploinsufficiency of *KDR* (encoding VEGF receptor 2) and, likely to a lesser extent *IQGAP1* (VEGF receptor signaling), contribute to TOF risk. Rare *KDR* loss-of-function variants were previously reported in several cohorts with TOF and other conotruncal defects ^18, 21, 30^ (Figure S1), and results from the current study of the largest TOF cohort available indicate a significant enrichment compared to controls (FET: *p* = 1.8E-4). Other studies report evidence that similar *KDR* variants are also risk factors for pulmonary arterial hypertension, independent of any heart malformations ^40, 41^. As for many CHD genes, this suggests pleiotropy for *KDR* requiring further study. There may also be clinical implications with respect to pulmonary hypertension in TOF ^42^. *IQGAP1* loss-of-function variants were identified in this and various other CHD cohorts ^18, 31, 32^ (including multiple *de novo* variants), but were also found in some parents of probands with autism. Besides its essential role in VEGF receptor signaling, IQGAP1 regulates and integrates other cellular processes, including neuronal functions ^43^. We consider *IQGAP1* a promising candidate gene for CHD, but further statistical support and phenotypic characterization will be needed.

Pathogenic/likely pathogenic and candidate variants were identified in a total of 26 genes: *ARHGAP31, ASXL1, ATRX, CACNA1C, CHD7, CSNK2A1, DLL4, EP300, FLT4, GATAD2B, GATA6, GDF1, JAG1, IQGAP1, KAT6A, KDR, LZTR1, NF1, NODAL, NOTCH1, PIK3CA, PSMD12, RAF1, RASA1, SMAD2*, and *TBX1*. The results further indicated that the encoded proteins form highly inter-connected networks of functional interaction. This suggests that the genetic heterogeneity identified through human disease studies may help to inform overlapping or unifying molecular pathomechanisms for TOF. Notably, VEGFR2 (*KDR*) and NOTCH1 form central nodes in this interaction network, supporting and extending evidence that the developing right outflow tract is vulnerable to VEGF/Notch dysregulation ^44-46^. VEGF signaling was also found as the top canonical pathway associated with *de novo* variants in conotruncal defects ^32^, and low VEGF expression was linked to TOF risk ^47^. Delineating the relevant protein networks and associated pathomechanisms will help to rank novel candidate genes and could inform potential therapeutic targets. For example, loss-of-function variants in *TCF12* (Table S4) were recently reported in multiple individuals with unexplained CHD ^30^, and the encoded protein functionally interacts with three confirmed TOF-associated proteins (NOTCH1, SMAD2, EP300).

Even with overlapping molecular functions, however, the phenotypic spectrum (pleiotropy) can vary largely not only from one gene to another, and one variant to another, but for individuals with the same variant within and between families. Most of the identified pathogenic/likely pathogenic variants in CHD genes were associated with multisystemic genetic disorders (Table S2), as expected for confirmed genes in early stages of clinical interpretation. Clinical genetic testing results may thus, in certain cases, flag the potential for an increased risk for comorbidities including neurodevelopmental delays. On the other hand, pathogenic variants historically identified through syndromic phenotypes may have cardiac phenotypes without classic extracardiac expression ^14^. For most individual genetic predispositions however, the extent of the disease spectrum is yet unknown. Delineating genotype-associated clinical traits, and understanding their penetrance, will be essential for genetic counselling, familial risk assessments, and informing outcome.

Identifying the genetic etiologies of TOF can improve clinical management, by providing information on outcomes and risks related to the variant, in addition to those related to the cardiac anatomical severity and other clinical parameters ^13, 48, 49^. After the surgical repair of TOF, heart failure and arrhythmias are leading causes of morbidity, impaired quality of life, and mortality. Genetic factors that can affect the molecular and structural properties of the heart and vasculature may play a role. In this study, 16 out of 811 probands (2.0%) were identified to have pathogenic/likely pathogenic variants that could affect cardiac surveillance and management recommendations (Table S7). Longitudinal clinical data will be needed to characterize adverse or favourable outcomes of patient populations that include such genetic variant data, in order to identify predictive markers and to inform preventive and therapeutic interventions, as part of precision medicine.

Advantages and limitations: We analyzed genetic risk variants in the largest available exome dataset of individuals with TOF. We prioritized variants with sufficient evidence for pathogenicity, according to consensus clinical guidelines ^25^, in order to increase the “signal-to-noise ratio” in the reporting of single gene defects. In contrast to primarily statistical approaches, such as that previously applied to this cohort ^20^, this clinical strategy helped enable us to take into account the expected genetic and allelic heterogeneity of pathogenic variants in TOF. For example, 18 genes were identified with only one pathogenic/likely pathogenic variant in this cohort. The lack of variant segregation data and the inaccessibility of individual clinical information, such as anatomical subtypes, disease progression, associated features, or age for this cohort ^20^, however limited the interpretation of the findings. Our analyses were also restricted to rare small sequence variants and insertions/deletions in regions targeted by exome sequencing, typically involving ∼95% of exonic regions. Despite their established contribution to CHD ^14^, the exome data available did not allow us to assess structural aberrations, such as rare copy number variants; individuals with typical 22q11.2 deletions were however excluded ^20^. All variants reported here were identified in heterozygous state, passed internal quality metrics, and were visually validated in the aligned sequencing reads, however we could not confirm their accuracy by Sanger sequencing.

## Conclusions

We studied the largest published exome sequencing dataset of patients with TOF, interpreting variants from the perspective of clinical pathogenicity. The identified genetic results add evidence for a major contribution of VEGF/Notch dysregulation and provide novel findings for functionally interacting protein networks relevant to the pathomechanism of TOF. We anticipate that clinical genomic sequencing, especially where capability of detecting structural variants is included, will become an essential component for assessing risks and outcomes in patients with CHD ^22^. Re-analysis of existing datasets is warranted, as our knowledge to identify and interpret disease-related variants continuously evolves.

## Data Availability

https://www.ebi.ac.uk/ega, study accession EGAS00001003302

## Acknowledgments

Sequence data has been accessed through the European Genome-phenome Archive (EGA), which is hosted by the EBI and the CRG, under accession number EGAS00001003302 (https://ega-archive.org). We thank Page et al.^20^ for allowing us access to the data, and the patients and their families for participation in this research effort. We thank the staff at The Centre for Applied Genomics (TCAG), a node of CGEn, for support in data analysis.

