## Supplemental material for "Clinical genetic risk variants inform a functional protein interaction network for tetralogy of Fallot"

### Supplementary methods

#### Exome sequencing data

We analyzed the exome sequencing data of a multi-centre cohort of children and adults with tetralogy of Fallot (TOF). Exome sequencing had been performed at the McGill University/Genome Quebec Innovation Centre (MUGQIC; Montreal, Canada), and the methodology is described in detail in the supplementary information of Page et al.[1]. Briefly, exome capture was performed using Agilent SureSelectXT Human All Exon 50 Mb version 4 kit (Agilent). Trimming of paired-end sequencing reads (100 bp), using Trimmomatic, and read mapping with removal of duplicate reads (BWA-MEM algorithm[2]) were done at the McGill Genome Centre.

We downloaded 829 bam files from the European Genome-phenome Archive (https://www.ebi.ac.uk/ega; EGAS00001003302)[1]. We excluded 18 data files that were: corrupted (n = 2), aligned to a different reference genome: bwa VN:0.7.6a-r433, b37 (n = 9), or found to be identical to other samples in the cohort (n = 7). We studied the resulting n = 811 unique exome sequencing datasets, aligned to the bwa VN:0.7.4-r385, hg19 reference genome.

#### Variant calling and annotation

The genome analysis tool kit (GATK v3.7; GATK Best Practices recommendations[3, 4]) was used for base quality score recalibration and indel realignment prior to variant calling using the HaplotypeCaller. Hard-filters were applied to the small nucleotide variants (SNVs) (QD < 2.0 || FS > 60.0 || MQ < 40.0 || MQRankSum < -12.5 || ReadPosRankSum< -8.0 || SOR > 3.0) and insertions/deletions (indels) (QD < 2.0 || FS > 200.0 || ReadPosRankSum< -20.0 || SOR > 10.0). Variant calls were annotated using a custom pipeline developed at TCAG based on ANNOVAR[5].

#### Variant assessments

We analyzed the exome sequencing dataset for rare (minor allele frequencies <0.1%) sequence variants (substitutions and small insertions/deletions), prioritizing genes associated with congenital heart disease (http://chdgene.victorchang.edu.au/), other cardiac disorders, or multi-systemic disorders (https://www.omim.org/, https://research.nhgri.nih.gov/CGD/).

(i) Predicted loss-of-function alleles

We predicted the following as loss-of-function (LoF) or null alleles: frameshift insertions, deletions or substitutions; substitutions creating a premature stop codon; and alterations of the intronic dinucleotide adjacent to a coding-exonic splice junction.

(ii) Allele frequency and control databases

Overall and population-specific allele frequencies for SNVs and small indels were derived from 1000 Genomes (African, American, East Asian, European, South Asian; http://www.internationalgenome.org/), ExAC (African, American, East Asian, Finnish, Non-Finnish Europeans, South Asians, Others; http://exac.broadinstitute.org/)[6], and gnomAD (African, American, Ashkenazi Jewish, East Asian, Finnish, Non-Finnish Europeans, South Asians, Others; http://gnomad.broadinstitute.org/)[7].

(iii) Disease gene and variant databases

Online Mendelian Inheritance in Man (OMIM; https://www.omim.org/) and CHDgene (http://chdgene.victorchang.edu.au/) were used as disease gene databases. The Human Gene Mutation Database (HGMD; http://www.hgmd.cf.ac.uk/ac/index.php) and ClinVar (https://www.ncbi.nlm.nih.gov/clinvar/) were used as disease variant databases.

(iv) Emerging candidate genes

We also analyzed for other genes with emerging evidence that they are associated with TOF or other congenital heart defects (https://pubmed.ncbi.nlm.nih.gov/). We assessed the available evidence in the literature, in combination with statistical support from the cohort reported here.

(v) Validation of variants

We only report on variants that were covered by read depths of ≥10x. Read alignments for all putative disease-associated variants were manually inspected using IGV (http://software.broadinstitute.org/software/igv/).

**Pathway enrichment analyses**

Genes annotated to ontology terms[8] were extracted using the Bioconductor R package GO.db v 3.5, while the pathways were built starting from their respective websites (for Reactome[9] and KEGG[10]), and Broad Institute website (for Biocarta) to create a custom gene-set collection. Gene-sets were filtered to retain gene ontology terms with a number of annotated genes between 15 and 1000, while Reactome, KEGG and Biocarta were filtered to retain only pathways with a number of annotated genes between 5 and 500. One-tailed Fisher exact test (Ha > H0) was used to calculate the enrichment p values, using all genes in the filtered collection as the universe. P values and odd ratios are reported in Table S8 (provided as a separate file); the Benjamini-Hochberg procedure was used for the multiple test comparison correction.

**Table S1. Exome sequencing coverage statistics.**

| Included datasets (n = 811) | Mean target coverage per exome | Proportion with coverage ≥ 10x |
|---|---|---|
| **Average** | 107.9x | 98.4% |
| **Median** | 111.0x | 98.6% |
| **Range** | 33.4x - 158.6x | 90.8% - 99.6% |

### Supplementary results

**Table S2. Pathogenic/likely pathogenic risk variants for congenital heart defects (CHD), identified in a cohort of 811 probands with tetralogy of Fallot (n = 40 loss-of-function, n = 10 missense in 23 genes; results summarized in Figure 1).**

| Sample | Gene | Variant | Variant type | MAF | Disease mechanism/evidence | OMIM-P # | OMIM disease (inheritance) or supporting literature |
|---|---|---|---|---|---|---|---|
| B009FTV[a] | *FLT4* (NM_182925.4) | c.2758C>T, p.(Gln920*) | LOF | 4.18E-06 | FLT4 haploinsufficiency | 618780 | Congenital heart defects, multiple types (AD) |
| B009FV1_R[a] | *FLT4* (NM_182925.4) | c.2300-1G>C, p.? | LOF | 0 | FLT4 haploinsufficiency | 618780 | Congenital heart defects, multiple types (AD) |
| B00B0EY_R[a] | *FLT4* (NM_182925.4) | c.1267dupC, p.(Gln423Profs*4) | LOF | 4.11E-06 | FLT4 haploinsufficiency | 618780 | Congenital heart defects, multiple types (AD) |
| B00B0HD[a] | *FLT4* (NM_182925.4) | c.2849_2850+18del CGCAGGCCGCCCGCTCACCG, p.? | LOF | 0 | FLT4 haploinsufficiency | 618780 | Congenital heart defects, multiple types (AD) |
| B00B0KK[a] | *FLT4* (NM_182925.4) | c.1083C>A, p.(Tyr361*) | LOF | 0 | FLT4 haploinsufficiency | 618780 | Congenital heart defects, multiple types (AD) |
| B00B0O8[a] | *FLT4* (NM_182925.4) | c.2686G>T, p.(Glu896*) | LOF | 0 | FLT4 haploinsufficiency | 616589 | Congenital heart defects, multiple types (AD) |
| B00B0PK[a] | *FLT4* (NM_182925.4) | c.2714delA, p.(Asn905Thrfs*21) | LOF | 0 | FLT4 haploinsufficiency | 616589 | Congenital heart defects, multiple types (AD |
| B00B0QE_R[a] | *FLT4* (NM_182925.4) | c.2559dupC, p.(Gly854Argfs*21) | LOF | 0 | FLT4 haploinsufficiency | 616589 | Congenital heart defects, multiple types (AD) |
| B00B16G[a] | *FLT4* (NM_182925.4) | c.3002-1G>A, p.? | LOF | 0 | FLT4 haploinsufficiency | 616589 | Congenital heart defects, multiple types (AD) |
| B00BDV3_R[a] | *FLT4* (NM_182925.4) | c.3376C>T, p.(Gln1126*) | LOF | 0 | FLT4 haploinsufficiency | 616589 | Congenital heart defects, multiple types (AD) |
| B00BDV4[a] | *FLT4* (NM_182925.4) | c.3002-2A>G, p.? | LOF | 0 | FLT4 haploinsufficiency | 616589 | Congenital heart defects, multiple types (AD) |
| B00BET4[a] | *FLT4* (NM_182925.4) | c.1107C>G, p.(Tyr369*) | LOF | 0 | FLT4 haploinsufficiency | 616589 | Congenital heart defects, multiple types (AD) |
| B00BETB[a] | *FLT4* (NM_182925.4) | c.1902_1906dupCACGC, p.(Leu636Profs*5) | LOF | 0 | FLT4 haploinsufficiency | 616589 | Congenital heart defects, multiple types (AD) |
| B00DKZL_R[a] | *FLT4* (NM_182925.4) | c.3091C>T, p.(Arg1031*) | LOF | 0 | FLT4 haploinsufficiency | 616589 | Congenital heart defects, multiple types (AD) |
| B00B0K9 | *NOTCH1* (NM_017617.3) | c.1045delA, p.(Thr349Profs*282) | LOF | 2.07E-05 | NOTCH1 haploinsufficiency | 616028, 109730 | Adams-Oliver syndrome (AD)[b], Aortic valve disease (AD) |
| B00B0O5[a] | *NOTCH1* (NM_017617.3) | c.5385-2delA, p.? | LOF | 0 | NOTCH1 haploinsufficiency | 616028, 109730 | Adams-Oliver syndrome (AD)[b], Aortic valve disease (AD) |

| | | | | | | | |
|---|---|---|---|---|---|---|---|
| B00B0SE[a,d] | *NOTCH1* (NM_017617.3) | c.1342C>T, p.(Arg448*) | LOF | 0 | NOTCH1 haploinsufficiency | 616028, 109730 | Adams-Oliver syndrome (AD)[b], Aortic valve disease (AD) |
| B00B0SS[a] | *NOTCH1* (NM_017617.3) | c.344delG, p.(Gly115Alafs*8) | LOF | 0 | NOTCH1 haploinsufficiency | 616028, 109730 | Adams-Oliver syndrome (AD)[b], Aortic valve disease (AD) |
| B00BDT8[a] | *NOTCH1* (NM_017617.3) | c.3966delC, p.(Cys1322Trpfs*123) | LOF | 0 | NOTCH1 haploinsufficiency | 616028, 109730 | Adams-Oliver syndrome (AD)[b], Aortic valve disease (AD) |
| B00BEAA[a] | *NOTCH1* (NM_017617.3) | c.5197C>T, p.(Gln1733*) | LOF | 0 | NOTCH1 haploinsufficiency | 616028, 109730 | Adams-Oliver syndrome (AD)[b], Aortic valve disease (AD) |
| B00DLD1[a] | *NOTCH1* (NM_017617.3) | c.440delA, p.(Asn147Thrfs*130) | LOF | 0 | NOTCH1 haploinsufficiency | 616028, 109730 | Adams-Oliver syndrome (AD)[b], Aortic valve disease (AD) |
| B009FQG_R[a] | *NOTCH1* (NM_017617.3) | c.599G>T, p.(Gly200Val) | Missense | 0 | De novo in Page et al.[1], found in two unrelated individuals | 616028, 109730 | Adams-Oliver syndrome (AD)[b], Aortic valve disease (AD) |
| B00B0KM[a] | *NOTCH1* (NM_017617.3) | c.599G>T, p.(Gly200Val) | Missense | 0 | De novo in Page et al.[1], found in two unrelated individuals | 616028, 109730 | Adams-Oliver syndrome (AD)[b], Aortic valve disease (AD) |
| B009G03_R[a] | *NOTCH1* (NM_017617.3) | c.5624A>G, p.(Asn1875Ser) | Missense | 0 | De novo, reduced Notch signaling in Page et al.[1] | 616028, 109730 | Adams-Oliver syndrome (AD)[b], Aortic valve disease (AD) |
| B009G7N[a] | *NOTCH1* (NM_017617.3) | c.1820G>A, p.(Cys607Tyr) | Missense | 0 | Reduced Notch signaling in Page et al.[1], cysteine substitution in EGF domain | 616028, 109730 | Adams-Oliver syndrome (AD)[b], Aortic valve disease (AD) |
| B009FUJ_R | *JAG1* (NM_000214.2) | c.1984delG, p.(Ala662Profs*81) | LOF | 0 | JAG1 haploinsufficiency | 187500, 118450 | Tetralogy of Fallot (AD), Alagille syndrome (AD)[b] |
| B00B0GY | *JAG1* (NM_000214.2) | c.2639_2640delGT, p.(Cys880*) | LOF | 0 | JAG1 haploinsufficiency | 187500, 118450 | Tetralogy of Fallot (AD), Alagille syndrome (AD)[b] |
| B00B0IL | *JAG1* (NM_000214.2) | c.2372+2T>G, p.? | LOF | 0 | JAG1 haploinsufficiency | 187500, 118450 | Tetralogy of Fallot (AD), Alagille syndrome (AD)[b] |
| B00B0KQ | *TBX1* (NM_080646.1) | c.549delG, p.(Lys184Argfs*24) | LOF | 0 | TBX1 haploinsufficiency | 187500 | Tetralogy of Fallot (AD) |
| B00DL8V | *TBX1* (NM_080646.1) | c.908+1G>A, p.? | LOF | 0 | TBX1 haploinsufficiency | 187500 | Tetralogy of Fallot (AD) |
| B00B0RO | *GATA6* (NM_005257.6) | c.1502C>A, p.(Ser501*) | LOF | 0 | GATA6 haploinsufficiency | 187500, 600001 | Tetralogy of Fallot (AD), Pancreatic agenesis and congenital heart defects (AD)[b] |
| B00BEBP | *GATA6* (NM_005257.6) | c.1367G>A, p.(Arg456His) | Missense | 0 | De novo in Allen et al.[11] | 187500, 600001 | Tetralogy of Fallot (AD), Pancreatic agenesis and congenital heart defects (AD)[b] |
| B009FO4_R | *KAT6A* (NM_006766.3) | c.1506delT, p.(Asp503Ilefs*42) | LOF | 0 | KAT6A haploinsufficiency | 616268 | Arboleda-Tham syndrome (AD)[b] |
| B009FO7[c] | *PSMD12* (NM_002816.5) | c.430C>T, p.(Arg144*) | LOF | 0 | PSMD12 haploinsufficiency | 617516 | Stankiewicz-Isidor syndrome (AD)[b] |
| B009G6B | *CSNK2A1* (NM_177559.2) | c.131_132delAA, p.(Lys44Ilefs*5) | LOF | 0 | CSNK2A1 haploinsufficiency | 617062 | Okur-Chung neurodevelopmental syndrome (AD)[b] |
| B00B0KC | *ATRX* (NM_000489.5) | c.3736+1G>T, p.? | LOF | 0 | ATRX deficiency | 301040 | Alpha-thalassemia/mental retardation syndrome (XLD)[b] |

| | | | | | | | |
|---|---|---|---|---|---|---|---|
| B00BEAD_R | *NF1* (NM_000267.3) | c.5206-1G>C, p.? | LOF | 0 | NF1 haploinsufficiency | 162200 | Neurofibromatosis, type 1 (AD)[b] |
| B00DLD6 | *CHD7* (NM_017780.4) | c.7160C>G, p.(Ser2387*) | LOF | 0 | CHD7 haploinsufficiency | 214800 | CHARGE syndrome (AD)[b] |
| B009FO7[c] | *ASXL1* (NM_015338.6) | c.2728C>T, p.(Gln910*) | LOF | 0 | ASXL1 haploinsufficiency | 605039 | Bohring-Opitz syndrome (AD)[b] |
| B009G7S | *GATAD2B* (NM_020699.4) | c.520C>T, p.(Arg174*) | LOF | 0 | GATAD2B haploinsufficiency[12] | 615074 | GAND syndrome (AD)[b] |
| B009G79_R | *PIK3CA* (NM_006218.4) | c.2809_2810del, p.F937fs | LOF | 0 | PIK3CA haploinsufficiency[13] | 602501 | Megalencephaly-capillary malformation-polymicrogyria syndrome (postzygotic variants)[b] |
| B009G7Z | *RASA1* (NM_002890.2) | c.2150_2151delTC, p.(Ile717Asnfs*8) | LOF | 0 | RASA1 haploinsufficiency[14] | 608354 | Capillary malformation-arteriovenous malformation (AD)[b] |
| B00B0IX | *NODAL* (NM_018055.5) | c.692G>A, p.(Trp231*) | LOF | 4.06E-06 | NODAL haploinsufficiency[15, 16] | 270100 | Heterotaxy (AD)[b] |
| B00DKWE | *ARHGAP31* (NM_020754.4) | c.2047C>T, p.(Gln683*) | LOF | 0 | ARHGAP31 haploinsufficiency | 100300 | Adams-Oliver syndrome (AD)[b] |
| B00DKW1 | *SMAD2* (NM_005901.6) | c.998-2A>C, p.? | LOF | 0 | SMAD2 haploinsufficiency | NA | [17, 18, b] |
| B00B0S0 | *DLL4* (NM_019074.4) | c.949A>C, p.(Thr317Pro) | Missense | 0 | De novo in Meester et al.[19] | 616589 | Adams-Oliver syndrome (AD)[b] |
| B009FV4 | *RAF1* (NM_001354689.3) | c.1532C>T, p.(Thr511Ile) | Missense | 0 | RAF1 gain-of-function, curated by ClinVar expert panel | 611553 | Noonan syndrome (AD)[b] |
| B009FVM_R | *CACNA1C* (NM_001167625.1) | c.1216G>A, p.(Gly406Arg) | Missense | 0 | De novo in Napolitano et al.[20] | 601005 | Timothy syndrome (AD)[b] |
| B00B0LX | *LZTR1* (NM_006767.4) | c.742G>A, p.(Gly248Arg) | Missense | 0 | De novo or segregating in Chinton et al.[21] | 616564 | Noonan syndrome (AD)[b] |
| B00BDTA | *EP300* (NM_001429.4) | c.4783T>G, p.(Phe1595Val) | Missense | 0 | De novo in Retterer et al.[22] | 613684 | Rubinstein-Taybi syndrome (AD)[b] |

All variants were heterozygous. Variant counts as displayed in Figure 1 (confirmed CHD genes). Minor allele frequencies (MAF) were derived from gnomAD_exomes_All.

[a] Variant also reported in Page et al.[1]. Note: Reporting of *NOTCH1*, *FLT4* and other variants may differ on the basis of differences in methodology of the current study to that of Page et al. [1], e.g., nomenclature, frequency cut-offs, in silico prediction methods, exclusion of samples (see discussion in manuscript text).

[b] Multi-systemic, or potentially multi-systemic condition.

[c] Individual B009FO7 had two likely pathogenic variants, one in *PSMD12* and one in *ASXL1*, both associated with an increased risk for CHD.

[d] Individual B00B0SE had two likely pathogenic/pathogenic variants, one in *NOTCH1* and one in *MYBPC3* with potential implications for cardiovascular outcome (Table S7).

AD, autosomal dominant; LOF, loss-of-function; NA, Not available; XLD, X-linked dominant.

**Table S3. Loss-of-function variants in emerging TOF/CHD candidate genes, identified in three or more individuals in the cohort studied (and with statistical support, as presented in Figure 1 and Results).**

| Sample | Gene | Variant | Variant type | MAF | Disease mechanism/evidence | OMIM-P # | OMIM disease (inheritance) or supporting literature |
|---|---|---|---|---|---|---|---|
| B009FP4 | *KDR* (NM_002253.2) | c.3580_3581delCT, p.(Leu1194Alafs*15) | LOF | 0 | KDR haploinsufficiency | NA | [23, 24] |
| B00B0PI_R | *KDR* (NM_002253.2) | c.2605delA, p.(Met869Cysfs*2) | LOF | 0 | KDR haploinsufficiency | NA | [23, 24] |
| B00BQTD | *KDR* (NM_002253.2) | c.3170delC, p.(Pro1057Glnfs*13) | LOF | 0 | KDR haploinsufficiency | NA | [23, 24] |
| B00DKWY | *KDR* (NM_002253.2) | c.2373+1G>A, p.? | LOF | 0 | KDR haploinsufficiency | NA | [23, 24] |
| B009FQA_R | *IQGAP1* (NM_003870.3) | c.650-2A>T, p.? | LOF | 1.33E-05 | IQGAP1 haploinsufficiency | NA | [23, 25, 26] |
| B00B1D6 | *IQGAP1* (NM_003870.3) | c.1393dupA, p.(Arg465Lysfs*26) | LOF | 0 | IQGAP1 haploinsufficiency | NA | [23, 25, 26] |
| B009FOS_R | *IQGAP1* (NM_003870.3) | c.1150C>T, p.(Gln384*) | LOF | 4.15E-06 | IQGAP1 haploinsufficiency | NA | [23, 25, 26] |
| B009FNV | *GDF1* (NM_001492.4) | c.681C>A, p.(Cys227*) | LOF | 9.00E-04[a] | GDF1 haploinsufficiency[27] | 613854 | Congenital heart defects, multiple types (AD) |
| B009FTS_R | *GDF1* (NM_001492.4) | c.681C>A, p.(Cys227*) | LOF | 9.00E-04[a] | GDF1 haploinsufficiency[27] | 613854 | Congenital heart defects, multiple types (AD) |
| B00B0H7 | *GDF1* (NM_001492.4) | c.681C>A, p.(Cys227*) | LOF | 9.00E-04[a] | GDF1 haploinsufficiency[27] | 613854 | Congenital heart defects, multiple types (AD) |
| B00B0JI | *GDF1* (NM_001492.4) | c.681C>A, p.(Cys227*) | LOF | 9.00E-04[a] | GDF1 haploinsufficiency[27] | 613854 | Congenital heart defects, multiple types (AD) |
| B00BDT4 | *GDF1* (NM_001492.4) | c.681C>A, p.(Cys227*) | LOF | 9.00E-04[a] | GDF1 haploinsufficiency[27] | 613854 | Congenital heart defects, multiple types (AD) |
| B00BDTU | *GDF1* (NM_001492.4) | c.681C>A, p.(Cys227*) | LOF | 9.00E-04[a] | GDF1 haploinsufficiency[27] | 613854 | Congenital heart defects, multiple types (AD) |
| B00DL0L | *GDF1* (NM_001492.4) | c.681C>A, p.(Cys227*) | LOF | 9.00E-04[a] | GDF1 haploinsufficiency[27] | 613854 | Congenital heart defects, multiple types (AD) |
| B009G0C | *GDF1* (NM_001492.4) | c.1047_1050delCTTT, p.(Phe349Leufs*35) | LOF | 1.00E-04[a] | GDF1 haploinsufficiency[27] | 613854 | Congenital heart defects, multiple types (AD) |

All variants were heterozygous. Variant counts as displayed in Figure 1 (candidate genes).

[a] Minor allele frequencies (MAF) for *GDF1* were derived from gnomAD_genomes_Non-Finnish Europeans. A comprehensive assessment of the *GDF1* locus could not be performed due to insufficient coverage (defined as read depths <10x; supplementary methods). Other MAF were derived from gnomAD_exomes_All.

LOF, loss-of-function; NA, Not available.

**Table S4. Pathogenic/likely pathogenic and candidate loss of function variants (n = 17) in 8 genes observed in a published independent cohort of 424 probands with tetralogy of Fallot (Jin et al. [28]).**

| Sample | Gene | Variant | MAF | Disease mechanism/evidence | OMIM-P # | OMIM disease (inheritance) or supporting literature |
|---|---|---|---|---|---|---|
| 1-00645 | *FLT4* (NM_182925.4) | c.2206C>T, p.(Gln736*) | 0 | FLT4 haploinsufficiency | 618780 | Congenital heart defects, multiple types (AD) |
| 1-01795 | *FLT4* (NM_182925.4) | c.1088dupC, p.(Pro364Alafs*63) | 0 | FLT4 haploinsufficiency | 618780 | Congenital heart defects, multiple types (AD) |
| 1-00788 | *FLT4* (NM_182925.4) | c.503_506delCGCT, p.(Thr168Serfs*76) | 0 | FLT4 haploinsufficiency | 618780 | Congenital heart defects, multiple types (AD) |
| 1-05967 | *FLT4* (NM_182925.4) | c.89delC, p.(Pro30Argfs*3) | 4.05E-05 | FLT4 haploinsufficiency | 618780 | Congenital heart defects, multiple types (AD) |
| 1-04970 | *FLT4* (NM_182925.4) | c.1083C>G, p.(Tyr361*) | 0 | FLT4 haploinsufficiency | 618780 | Congenital heart defects, multiple types (AD) |
| 1-06642 | *FLT4* (NM_182925.4) | c.2804delT, p.(Leu935Profs*72) | 0 | FLT4 haploinsufficiency | 618780 | Congenital heart defects, multiple types (AD) |
| 1-03410 | *FLT4* (NM_182925.4) | c.89delC, p.(Pro30Argfs*3) | 4.05E-05 | FLT4 haploinsufficiency, de novo | 618780 | Congenital heart defects, multiple types (AD) |
| 1-03825 | *FLT4* (NM_182925.4) | c.2844_2845delCT, p.(Cys949Argfs*53) | 0 | FLT4 haploinsufficiency, de novo | 618780 | Congenital heart defects, multiple types (AD) |
| 1-00305 | *NOTCH1* (NM_017617.3) | c.273_277delGGGCT, p.(Gly92Leufs*49) | 0 | NOTCH1 haploinsufficiency | 616028, 109730 | Adams-Oliver syndrome (AD)[d], Aortic valve disease (AD) |
| 1-00692 | *NOTCH1* (NM_017617.3) | c.1800_1801dupCG, p.(Glu601Alafs*31) | 0 | NOTCH1 haploinsufficiency | 616028, 109730 | Adams-Oliver syndrome (AD)[d], Aortic valve disease (AD) |
| 1-02338 | *NOTCH2* (NM_024408.3) | c.4299delA, p.(Ala1434Leufs*119) | 0 | NOTCH2 haploinsufficiency | 610205 | Alagille syndrome (AD) |
| 1-00534 | *CHD7* (NM_017780.3) | c.4795C>T, p.(Gln1599*) | 0 | CHD7 haploinsufficiency, de novo | 214800 | CHARGE syndrome (AD) |
| 1-08360 | *CHD7* (NM_017780.3) | c.4393C>T, p.(Arg1465*) | 0 | CHD7 haploinsufficiency, de novo | 214800 | CHARGE syndrome (AD) |
| 1-04135 | *MEIS2* (NM_170675.4) | c.383delA, p.(Lys128Serfs*19) | 0 | MEIS2 haploinsufficiency, de novo | 600987 | Cleft palate, cardiac defects, and mental retardation (AD) |
| 1-00141 | *NAA15* (NM_057175.3) | c.2282C>A, p.(Ser761*) | 0 | NAA15 haploinsufficiency, de novo | 617787 | Mental retardation (AD) |
| 1-08084 | *RPL5* (NM_000969.3) | c.67C>T, p.(Arg23*) | 0 | RPL5 haploinsufficiency, de novo | 612561 | Diamond-Blackfan anemia (AD) |
| 1-07375 | *KDR* (NM_002253.2) | c.1585A>T, p.(Lys529*) | 0 | KDR haploinsufficiency | NA | [23, 24] |

Variants were extracted from variant lists in Jin et al., supplementary Tables S7 and S9[28], and interpreted according to consensus guidelines[29]. This approach resulted in a lower yield, compared to the re-analysis of raw exome files from Page et al.[1] (Tables S2 and S3). Minor allele frequencies (MAF) were derived from gnomAD_exomes_All.

NA, not available.

**Table S5. Additional rare variants, meeting criteria for predicted deleterious variants of uncertain significance (n = 24 loss-of-function, n = 47 missense, n = 9 other) in CHD-relevant genes (n = 19) or candidate genes (n = 8).**

| Sample | Gene | Variant | Variant type | MAF | Annotation | OMIM-P # | OMIM disease (inheritance) or supporting literature |
|---|---|---|---|---|---|---|---|
| **VUS in CHD-relevant genes (OMIM, http://chdgene.victorchang.edu.au/)** | | | | | | | |
| B00DLES[a] | *FLT4* (NM_182925.4) | c.4011delT, p.(Tyr1337*) | Other VUS (premature stopcodon in the last exon) | 0 | NA | 618780 | Congenital heart defects, multiple types (AD) |
| B00B0KK[a,b] | *FLT4* (NM_182925.4) | c.781G>T, p.(Gly261Cys) | Missense VUS | 0 | Predicted damaging (CADD 24.3) | 618780 | Congenital heart defects, multiple types (AD) |
| B00B0LF[a] | *FLT4* (NM_182925.4) | c.153C>G, p.(Cys51Trp) | Missense VUS | 0 | Predicted damaging (CADD 34) | 618780 | Congenital heart defects, multiple types (AD) |
| B00BDS0 | *FLT4* (NM_182925.4) | c.513G>A, p.(Ser171Ser) | Other VUS (synonymous) | 0 | Predicted splice effect (CADD 20.5) | 618780 | Congenital heart defects, multiple types (AD) |
| B00BDS9[a] | *FLT4* (NM_182925.4) | c.89C>T, p.(Pro30Leu) | Missense VUS | 0 | Predicted damaging (CADD 25.6) | 618780 | Congenital heart defects, multiple types (AD) |
| B00BDSV[a] | *FLT4* (NM_182925.4) | c.89C>G, p.(Pro30Arg) | Missense VUS | 0 | Predicted damaging (CADD 24.9) | 618780 | Congenital heart defects, multiple types (AD) |
| B00BECK[a] | *FLT4* (NM_182925.4) | c.482C>T, p.(Ser161Phe) | Missense VUS | 0 | Predicted damaging (CADD 25.4) | 618780 | Congenital heart defects, multiple types (AD) |
| B00DL8L[a] | *FLT4* (NM_182925.4) | c.2284A>G, p.(Ser762Gly) | Missense VUS | 0 | Predicted damaging (CADD 24.1) | 618780 | Congenital heart defects, multiple types (AD) |
| B009FTP[a] | *NOTCH1* (NM_017617.3) | c.4646G>A, p.(Cys1549Tyr) | Missense VUS | 0 | De novo in Page et al.[1] | 616028, 109730 | Adams-Oliver syndrome (AD), Aortic valve disease (AD) |
| B009FNG_R[a] | *NOTCH1* (NM_017617.3) | c.428C>T, p.(Pro143Leu) | Missense VUS | 0 | Predicted damaging (CADD 24.7), in three probands | 616028, 109730 | Adams-Oliver syndrome (AD), Aortic valve disease (AD) |
| B00BDSY_R[a] | *NOTCH1* (NM_017617.3) | c.428C>T, p.(Pro143Leu) | Missense VUS | 0 | Predicted damaging (CADD 24.7), in three probands | 616028, 109730 | Adams-Oliver syndrome (AD), Aortic valve disease (AD) |
| B00BDTA[a,b] | *NOTCH1* (NM_017617.3) | c.428C>T, p.(Pro143Leu) | Missense VUS | 0 | Predicted damaging (CADD 24.7), in three probands | 616028, 109730 | Adams-Oliver syndrome (AD), Aortic valve disease (AD) |
| B009G8F[a] | *NOTCH1* (NM_017617.3) | c.1057C>T, p.(Arg353Cys) | Missense VUS | 0 | Predicted damaging (CADD 33), in two probands | 616028, 109730 | Adams-Oliver syndrome (AD), Aortic valve disease (AD) |
| B00B16Z[a] | *NOTCH1* (NM_017617.3) | c.1057C>T, p.(Arg353Cys) | Missense VUS | 0 | Predicted damaging (CADD 33), in two probands | 616028, 109730 | Adams-Oliver syndrome (AD), Aortic valve disease (AD) |

| | | | | | | | |
|---|---|---|---|---|---|---|---|
| B00B0D6[a] | *NOTCH1* (NM_017617.3) | c.5497G>C, p.(Asp1833His) | Missense VUS | 0 | Predicted damaging (CADD 24.3) | 616028, 109730 | Adams-Oliver syndrome (AD), Aortic valve disease (AD) |
| B00B0IB | *NOTCH1* (NM_017617.3) | c.1687T>C, p.(Cys563Arg) | Missense VUS | 0 | Predicted damaging (CADD 25.3) | 616028, 109730 | Adams-Oliver syndrome (AD), Aortic valve disease (AD) |
| B00B0IH[a] | *NOTCH1* (NM_017617.3) | c.875G>A, p.(Cys292Tyr) | Missense VUS | 0 | Predicted damaging (CADD 25.9) | 616028, 109730 | Adams-Oliver syndrome (AD), Aortic valve disease (AD) |
| B00B0IW[a] | *NOTCH1* (NM_017617.3) | c.6011G>T, p.(Arg2004Leu) | Missense VUS | 0 | Predicted damaging (CADD 33) | 616028, 109730 | Adams-Oliver syndrome (AD), Aortic valve disease (AD) |
| B00B0KE[a] | *NOTCH1* (NM_017617.3) | c.545G>A, p.(Cys182Tyr) | Missense VUS | 0 | Predicted damaging (CADD 27.6) | 616028, 109730 | Adams-Oliver syndrome (AD), Aortic valve disease (AD) |
| B00B0KI[a] | *NOTCH1* (NM_017617.3) | c.5017G>C, p.(Gly1673Arg) | Missense VUS | 0 | Predicted damaging (CADD 28.2) | 616028, 109730 | Adams-Oliver syndrome (AD), Aortic valve disease (AD) |
| B00B0KR[a] | *NOTCH1* (NM_017617.3) | c.4428_4430delCGG, p.(Gly1477del) | Other VUS (in-frame deletion) | 0 | NA | 616028, 109730 | Adams-Oliver syndrome (AD), Aortic valve disease (AD) |
| B00B0M0[a] | *NOTCH1* (NM_017617.3) | c.4483C>A, p.(Gln1495Lys) | Missense VUS | 0 | Predicted damaging (CADD 23.2) | 616028, 109730 | Adams-Oliver syndrome (AD), Aortic valve disease (AD) |
| B00B0SN[a] | *NOTCH1* (NM_017617.3) | c.1412T>A, p.(Ile471Asn) | Missense VUS | 0 | Predicted damaging (CADD 25.4) | 616028, 109730 | Adams-Oliver syndrome (AD), Aortic valve disease (AD) |
| B00B16Y[a] | *NOTCH1* (NM_017617.3) | c.1934G>A, p.(Cys645Tyr) | Missense VUS | 0 | Predicted damaging (CADD 27.6) | 616028, 109730 | Adams-Oliver syndrome (AD), Aortic valve disease (AD) |
| B00B187[a] | *NOTCH1* (NM_017617.3) | c.3974C>T, p.(Ala1325Val) | Missense VUS | 0 | Predicted damaging (CADD 25.2) | 616028, 109730 | Adams-Oliver syndrome (AD), Aortic valve disease (AD) |
| B00BEAA[a,b] | *NOTCH1* (NM_017617.3) | c.3880G>A, p.(Glu1294Lys) | Missense VUS | 0 | Predicted damaging (CADD 25.2) | 616028, 109730 | Adams-Oliver syndrome (AD), Aortic valve disease (AD) |
| B00BEAD_R[a,b] | *NOTCH1* (NM_017617.3) | c.598G>C, p.(Gly200Arg) | Missense VUS | 0 | Predicted damaging (CADD 28.6) | 616028, 109730 | Adams-Oliver syndrome (AD), Aortic valve disease (AD) |
| B00DKZ8[a] | *NOTCH1* (NM_017617.3) | c.490T>G, p.(Cys164Gly) | Missense VUS | 0 | Predicted damaging (CADD 26.8) | 616028, 109730 | Adams-Oliver syndrome (AD), Aortic valve disease (AD) |
| B00DKZI_R[a] | *NOTCH1* (NM_017617.3) | c.436_450dup TCCAACCCCTGCGCC, p.(Ser146_Ala150dup) | Other VUS (in-frame duplication) | 0 | NA | 616028, 109730 | Adams-Oliver syndrome (AD), Aortic valve disease (AD) |
| B00BDSL | *JAG1* (NM_000214.2) | c.185G>A, p.(Gly62Glu) | Missense VUS | 0 | Predicted damaging (CADD 22.5) | 187500, 118450 | Tetralogy of Fallot (AD), Alagille syndrome (AD) |
| B00BESY | *JAG1* (NM_000214.2) | c.376T>C, p.(Phe126Leu) | Missense VUS | 0 | Predicted damaging (CADD 33) | 187500, 118450 | Tetralogy of Fallot (AD), Alagille syndrome (AD) |
| B00BETA | *JAG1* (NM_000214.2) | c.2365C>T, p.(His789Tyr) | Missense VUS | 0 | Predicted damaging (CADD 23.3) | 187500, 118450 | Tetralogy of Fallot (AD), Alagille syndrome (AD) |
| B00B0SJ | *TBX1* (NM_080646.1) | c.1095_1101delAGGTGGC, p.(Gly366Valfs*2) | Other VUS (premature stopcodon in the last exon) | 0 | Does not affect all transcripts | 187500 | Tetralogy of Fallot (AD) |

| | | | | | | | |
|---|---|---|---|---|---|---|---|
| B00B0RM | *TBX1* (NM_080646.1) | c.607G>C, p.(Gly203Arg) | Missense VUS | 0 | Predicted damaging (CADD 31) | 187500 | Tetralogy of Fallot (AD) |
| B00BEST | *TBX1* (NM_080646.1) | c.840+5G>A, p.? | Other VUS (intronic) | 0 | Predicted splice effect (CADD 22.8) | 187500 | Tetralogy of Fallot (AD) |
| B00DKWP | *TBX1* (NM_080646.1) | c.956C>G, p.(Ala319Gly) | Missense VUS | 0 | Predicted damaging (CADD 28) | 187500 | Tetralogy of Fallot (AD) |
| B009FZL | *DLL4* (NM_019074.4) | c.1285C>T, p.(Arg429Cys) | Missense VUS | 0 | Predicted damaging (CADD 33), in three probands | 616589 | Adams-Oliver syndrome (AD) |
| B00BDSW | *DLL4* (NM_019074.4) | c.1285C>T, p.(Arg429Cys) | Missense VUS | 0 | Predicted damaging (CADD 33), in three probands | 616589 | Adams-Oliver syndrome (AD) |
| B00BDUG | *DLL4* (NM_019074.4) | c.1285C>T, p.(Arg429Cys) | Missense VUS | 0 | Predicted damaging (CADD 33), in three probands | 616589 | Adams-Oliver syndrome (AD) |
| B009FWD_R | *DLL4* (NM_019074.4) | c.1240G>A, p.(Gly414Arg) | Missense VUS | 0 | Predicted damaging (CADD 34) | 616589 | Adams-Oliver syndrome (AD) |
| B00BDUC | *CHD4* (NM_001273.2) | c.4515+1G>T, p.? | LOF VUS | 0 | Uncertain haplosensitivity of CHD4[30] | 617159 | Sifrim-Hitz-Weiss syndrome (AD) |
| B00B0H9 | *CHD4* (NM_001273.2) | c.2084G>A, p.(Arg695Gln) | Missense VUS | 0 | Predicted damaging (CADD 22.6) | 617159 | Sifrim-Hitz-Weiss syndrome (AD) |
| B00BDU2 | *CHD4* (NM_001273.2) | c.4513_4515delAAG, p.(Lys1505del) | Other VUS (in-frame deletion) | 0 | NA | 617159 | Sifrim-Hitz-Weiss syndrome (AD) |
| B00B0J2 | *ELN* (NM_000501.4) | c.1150+1G>A, p.? | Splice-site VUS | 5.69E-05 | ELN haploinsufficiency in aortic stenosis and other CHD[31] | 185500 | Supravalvar aortic stenosis (AD) |
| B009FVY_R | *ECE1* (NM_001397.3) | c.1021-2A>G, p.? | LOF VUS | 0 | Uncertain haplosensitivity of ECE1 | 613870 | Hirschsprung disease, cardiac defects, and autonomic dysfunction (AD) |
| B009G7T | *CACNA1C* (NM_001167625.1) | c.6169G>T, p.(Glu2057*) | LOF VUS | 0 | Uncertain haplosensitivity of CACNA1C | 601005 | Timothy syndrome (AD) |
| B009G10_R | *TLL1* (NM_001204760.1) | c.1159-2A>C, p.? | LOF VUS | 2.00E-04 | Uncertain haplosensitivity of TLL1 | 613087 | Atrial septal defect (AD) |
| B009FT1 | *TLL1* (NM_001204760.1) | c.713T>C, p.(Val238Ala) | Missense VUS | 2.00E-04 | Predicted damaging (CADD 22.7)[32], in two probands | 613087 | Atrial septal defect (AD) |
| B00B160_R | *TLL1* (NM_001204760.1) | c.713T>C, p.(Val238Ala) | Missense VUS | 2.00E-04 | Predicted damaging (CADD 22.7)[32], in two probands | 613087 | Atrial septal defect (AD) |

| | | | | | | | |
|---|---|---|---|---|---|---|---|
| B009G8X | *CRELD1* (NM_001077415.2) | c.959delA, p.(Gln320Argfs*25) | LOF VUS | 3.00E-04 | Uncertain haplosensitivity of CRELD1 | 606217 | Atrioventricular septal defect, partial, with heterotaxy syndrome (AD) |
| B00B0KJ[b] | *CRELD1* (NM_001077415.2) | c.977_984dupGCGGTTAT, p.(Arg329Alafs*19) | LOF VUS | 5.28E-05 | Uncertain haplosensitivity of CRELD1 | 606217 | Atrioventricular septal defect, partial, with heterotaxy syndrome (AD) |
| B00BDUD | *CRELD1* (NM_001077415.2) | c.484C>G, p.(Pro162Ala) | Missense VUS | 0 | Predicted damaging[33] | 606217 | Atrioventricular septal defect, partial, with heterotaxy syndrome (AD) |
| B00B0T7 | *SMAD6* (NM_005585.5) | c.223C>T, p.(Arg75*) | LOF VUS | 0 | Uncertain haplosensitivity of SMAD6 | 179300, 614823, 617439 | Radioulnar synostosis, nonsyndromic (AD), Aortic valve disease (AD), Craniosynostosis (AD) |
| B009FTM | *PRKD1* (NM_002742.2) | c.2241delC, p.(Ala748Leufs*8) | LOF VUS | 0 | Uncertain haplosensitivity of PRKD1 | 617364 | Congenital heart defects and ectodermal dysplasia (AD) |
| B009G10_R | *PRKD1* (NM_002742.2) | c.802A>T, p.(Lys268*) | LOF VUS | 2.44E-05 | Uncertain haplosensitivity of PRKD1 | 617364 | Congenital heart defects and ectodermal dysplasia (AD) |
| B00BET4 | *GLI3* (NM_000168.6) | c.2119C>T, p.(Pro707Ser) | Missense VUS | 3.00E-04 | Predicted damaging[34] | 146510 | Pallister-Hall syndrome (AD) |
| B00DKW1 | *KANSL1* (NM_001193466.2) | c.190C>T, p.(Arg64*) | Other VUS (premature stopcodon in the last exon) | 4.06E-06 | NA | 610443 | Koolen-De Vries syndrome (AD) |
| B009FR7_R | *MYH6* (NM_002471.3) | c.642+1G>T, p.? | LOF VUS | 0 | Uncertain haplosensitivity of MYH6 | 614089 | Atrial septal defect |
| B009FNJ_R | *NKX2-6* (NM_001136271.2) | c.797delG, p.(Gly266Valfs*?) | LOF VUS | 0 | Uncertain haplosensitivity of NKX2-6 | 217095 | Conotruncal heart malformations, Persistent truncus arteriosus |
| B009G3M_R | *NKX2-6* (NM_001136271.2) | c.274+1G>A, p.? | LOF VUS | 0 | Uncertain haplosensitivity of NKX2-6 | 217095 | Conotruncal heart malformations, Persistent truncus arteriosus |
| B009G8Y | *NKX2-6* (NM_001136271.2) | c.455dupA, p.(Gln153Alafs*?) | LOF VUS | 3.28E-05 | Uncertain haplosensitivity of NKX2-6 | 217095 | Conotruncal heart malformations, Persistent truncus arteriosus |
| B00B0LO | *NKX2-6* (NM_001136271.2) | c.455dupA, p.(Gln153Alafs*?) | LOF VUS | 3.28E-05 | Uncertain haplosensitivity of NKX2-6 | 217095 | Conotruncal heart malformations, Persistent truncus arteriosus |
| B00DL8V | *TAB2* (NM_001292034.3) | c.688C>A, p.(Gln230Lys) | Missense VUS | 1.63E-05 | Predicted damaging[35] | 614980 | Congenital heart defects, nonsyndromic (AD) |

| | | | | | | | |
|---|---|---|---|---|---|---|---|
| B00B0SL | *TBX20* (NM_001077653.2) | c.456C>G, p.(Ile152Met) | Missense VUS | 2.03E-05 | Predicted damaging[36] | 611363 | Atrial septal defect |
| **VUS in CHD candidate genes** | | | | | | | |
| B00B0KS | *KDR* (NM_002253.2) | c.3817G>T, p.(Glu1273*) | Other VUS (premature stopcodon in the penultimate exon) | 4.07E-06 | NA | NA | [23, 24] |
| B00B0RT_R | *KDR* (NM_002253.2) | c.2248G>C, p.(Ala750Pro) | Missense VUS | 0 | Predicted damaging (CADD 21.9) | NA | [23, 24] |
| B00BEB6 | *KDR* (NM_002253.2) | c.2234G>C, p.(Cys745Ser) | Missense VUS | 0 | Predicted damaging (CADD 25.4) | NA | [23, 24] |
| B00DL8L | *KDR* (NM_002253.2) | c.358G>C, p.(Asp120His) | Missense VUS | 0 | Predicted damaging (CADD 34) | NA | [23, 24] |
| B00B16V | *IQGAP1* (NM_003870.3) | c.641A>G, p.(Glu214Gly) | Missense VUS | 0 | Predicted damaging (CADD 28.8) | NA | [23, 25, 26] |
| B00BDSJ_R | *IQGAP1* (NM_003870.3) | c.3887A>G, p.(Tyr1296Cys) | Missense VUS | 0 | Predicted damaging (CADD 32) | NA | [23, 25, 26] |
| B00DLDA | *IQGAP1* (NM_003870.3) | c.353A>C, p.(Gln118Pro) | Missense VUS | 0 | Predicted damaging (CADD 24.6) | NA | [23, 25, 26] |
| B00BET7 | *VEGFA* (NM_001171623.1) | c.28delT, p.(Trp10Glyfs*32) | LOF VUS | 0 | VEGFA dysregulation in mouse models with TOF, candidate gene in humans with TOF | NA | [23,37] |
| B009FQG_R | *BCAR1* (NM_001170715.1) | c.1957C>T, p.(Arg653*) | LOF VUS | 0 | Candidate gene for TOF | NA | [23] |
| B009FWJ_R | *KIRREL3* (NM_032531.3) | c.634delC, p.(Leu212Serfs*105) | LOF VUS | 0 | In Jacobsen syndrome critical region | NA | NA |
| B009FUS_R | *LTBP3* (NM_001130144.2) | c.3376_3377delAG, p.(Ser1126Profs*85) | LOF VUS | 0 | Uncertain haplosensitivity of LTBP3, associated with cardiac defects[38] | 617809 | Geleophysic dysplasia (AD) |
| B00B0DN_R | *LTBP3* (NM_001130144.2) | c.1075delT, p.(Cys359Alafs*29) | LOF VUS | 4.06E-06 | Uncertain haplosensitivity of LTBP3, associated with cardiac defects[38] | 617809 | Geleophysic dysplasia (AD) |
| B00B0E9 | *ZFPM1* (NM_153813.2) | c.886delC, p.(Gln296Serfs*28) | LOF VUS | 0 | Candidate gene for TOF | NA | [39] |
| B00B0KF | *ZFPM1* (NM_153813.2) | c.1614dupC, p.(Ala539Argfs*133) | LOF VUS | 2.55E-05 | Candidate gene TOF | NA | [39] |
| B00B0NS_R | *MESP1* (NM_018670.3) | c.310G>T, p.(Glu104*) | LOF VUS | 6.28E-05 | Candidate gene for TOF | NA | [40, 41] |

| | | | | | | | |
|---|---|---|---|---|---|---|---|
| B00B16X | *MESP1* (NM_018670.3) | c.370G>T, p.(Glu124*) | LOF VUS | 4.99E-05 | Candidate gene for TOF | NA | [40, 41] |

All variants were heterozygous. Minor allele frequencies (MAF) were derived from gnomAD_exomes_All.

[a] Variant also reported in Page et al.[1]. Reporting of *NOTCH1*, *FLT4* and other variants may differ on the basis of differences in methodology of the current study to that of Page et al. [1], e.g., nomenclature, frequency cut-offs, in silico prediction methods, exclusion of samples (see Discussion in manuscript text).

[b] Individuals with other likely pathogenic variants (Table S2, S3).

LOF, loss-of-function; NA, Not available; VUS, variant of uncertain significance.

**Table S6. Other very rare, pathogenic/likely pathogenic variants in 7 genes for childhood-onset disorders (n = 8 loss-of-function, n = 1 missense), but with uncertain relevance currently to TOF.**

| Sample | Gene | Variant | Variant type | MAF | Disease mechanism/evidence | OMIM-P # | OMIM disease (inheritance) or supporting literature |
|---|---|---|---|---|---|---|---|
| B00DKWN | *TCF12*[b] (NM_207037.1) | c.556delG, p.(Val186Cysfs*59) | LOF | 0 | TCF12 haploinsufficiency | 615314 | Craniosynostosis (AD), [24] |
| B00B0KJ | *TCF12*[b] (NM_207037.1) | c.1640_1641delAA, p.(Lys547Argfs*4) | LOF | 0 | TCF12 haploinsufficiency | 615314 | Craniosynostosis (AD), [24] |
| B00B1CK | *POLR1A*[b] (NM_015425.6) | c.5062+1G>C, p.? | LOF | 0 | POLR1A haploinsufficiency | 616462 | Acrofacial dysostosis, Cincinnati type (AD), [42] |
| B00BEAR | *POLR1A*[b] (NM_015425.6) | c.3394C>T, p.(Arg1132*) | LOF | 0 | POLR1A haploinsufficiency | 616462 | Acrofacial dysostosis, Cincinnati type (AD), [42] |
| B00BDSE | *GLI2*[b] (NM_005270.5) | c.2293+1G>C, p.? | LOF | 0 | GLI2 haploinsufficiency | 615849, 610829 | Culler-Jones syndrome (AD), Holoprosencephaly (AD) |
| B00DL0I | *APC*[b] (NM_000038.6) | c.1213C>T, p.(Arg405*) | LOF | 0 | APC haploinsufficiency | 175100 | Adenomatous polyposis coli (AD) |
| B00B15G | *EDA* (NM_001399.5) | c.466C>T, p.(Arg156Cys) | Missense | 0 | De novo in Monreal et al.[43] | 313500 | Tooth agenesis, selective, X-linked 1 (XLD) |
| B00B0K7 | *ZMYND11* (NM_006624.5) | c.1203_1206delTCAA, p.(Asn401Lysfs*17) | LOF | 0 | ZMYND11 haploinsufficiency | 616083 | Mental retardation (AD) |
| B00BEAF[a] | *TRIO* (NM_007118.2) | c.3217dupG, p.(Ala1073Glyfs*34) | LOF | 0 | TRIO haploinsufficiency | 617061 | Intellectual developmental disorder, with microcephaly (AD) |

All variants were heterozygous. Minor allele frequencies (MAF) were derived from gnomAD_exomes_All.

[a] Individual with another likely pathogenic variant in *SCN5A* with potential implications for cardiovascular outcome (Table S7).

[b] Despite their uncertain relevance to TOF, the encoded proteins connect to the network map of TOF predisposing genes/proteins (Figure 2): GLI2 (NOTCH1, SMAD2, JAG1, PSMD12), APC (NOTCH1, IQGAP1, PSMD12, CSNK2A1), TCF12 (NOTCH1, SMAD2, EP300), POLR1A (EP300).

AD, autosomal dominant; LOF, loss-of-function; XLD, X-linked dominant.

**Table S7. Pathogenic/likely pathogenic variants (n = 16) with potential implications to cardiovascular outcome and management.**

| Sample | Gene | Variant | Variant interpretation | MAF | Disease | Cardiovascular risk | |
|---|---|---|---|---|---|---|---|
| | | | | | | CHD | Other outcomes |
| **Variants considered causative for the CHD (as presented in Table S2)** | | | | | | | |
| B00B0LX | *LZTR1* (NM_006767.4) | c.742G>A, p.(Gly248Arg) | Pathogenic | 0 | Noonan syndrome | TOF, pulmonary valve stenosis, septal defects, aortic coarctation, and other CHD[44] | Cardiac hypertrophy |
| B009FV4 | *RAF1* (NM_001354689.3) | c.1532C>T, p.(Thr511Ile) | Pathogenic | 0 | | | |
| B009FVM_R | *CACNA1C* (NM_001167625.1) | c.1216G>A, p.(Gly406Arg) | Pathogenic | 0 | Timothy syndrome | TOF, patent ductus arteriosus, patent foramen ovale, septal defects[20] | Arrhythmias, cardiac hypertrophy/dysfunction, sudden cardiac death |
| B00BEAD_R | *NF1* (NM_000267.3) | c.5206-1G>C, p.? | Likely pathogenic | 0 | Neurofibromatosis | TOF and other CHD | Cardiac hypertrophy, pulmonary hypertension, intracardiac neurofibroma, arterial hypertension, strokes |
| B009G7Z | *RASA1* (NM_002890.3) | c.2150_2151delTC, p.(Ile717Asnfs*8) | Likely pathogenic | 0 | Capillary malformation-arteriovenous malformation syndrome | TOF and other CHD[14] | Cardiac overload/heart failure due to arteriovenous malformations/fistulas |
| **Other cardiac risk variants** | | | | | | | |
| B00BDVA | *MYBPC3* (NM_000256.3) | c.1483C>G, p.(Arg495Gly) | Likely pathogenic | 4.06E-06 | Hypertrophic cardiomyopathy | Unknown | Cardiac hypertrophy, arrhythmias, heart failure, sudden cardiac death |
| B00B0SE[a] | *MYBPC3* (NM_000256.3) | c.1504C>T, p.(Arg502Trp) | Pathogenic | 4.87E-05 | | | |
| B00DLDV | *MYBPC3* (NM_000256.3) | c.3288delG, p.(Glu1096Aspfs*93) | Pathogenic | 0 | | | |
| B00BET1 | *MYH7* (NM_000257.4) | c.2594A>G, p.(Lys865Arg) | Likely pathogenic | 4.06E-06 | | | |
| B00B0HU | *MYL2* (NM_000432.4) | c. 64G>A, p.(Glu22Lys) | Likely pathogenic | 2.03E-05 | | | |
| B00BDUN | *TNNI3* (NM_000363.5) | c.484C>T, p.(Arg162Trp) | Likely pathogenic | 4.07E-05 | | | |
| B00B0JX | *DSC2* (NM_024422.6) | c.501_502delTA, p.(Thr168Hisfs*11) | Likely pathogenic | 0 | Arrhythmogenic right ventricular cardiomyopathy | Unknown | Arrhythmias, heart failure, sudden cardiac death |
| B00B178 | *DSP* (NM_004415.4) | c.7756C>T, p.(Arg2586*) | Likely pathogenic | 0 | | | |
| B00BDS5 | *DSP* (NM_004415.4) | c.8077_8080delAAAG, p.(Lys2693Profs*3) | Likely pathogenic | 0 | | | |
| B00B0D3 | *DMD* (NM_000109.4) | c.1789-1G>C, p.? | Likely pathogenic | 0 | Dilated cardiomyopathy | Unknown | Cardiac dilation, arrhythmias |

| | | | | | | | |
|---|---|---|---|---|---|---|---|
| B00BEAF[a] | *SCN5A* (NM_198056.2) | c.2692G>T, p.(Glu898*) | Likely pathogenic | 0 | Brugada syndrome | Unknown | Arrhythmias, sudden cardiac death |

All variants were heterozygous. Minor allele frequencies (MAF) were derived from gnomAD_exomes_All.

[a] Individuals with additional likely pathogenic variants (Tables S2, S6).

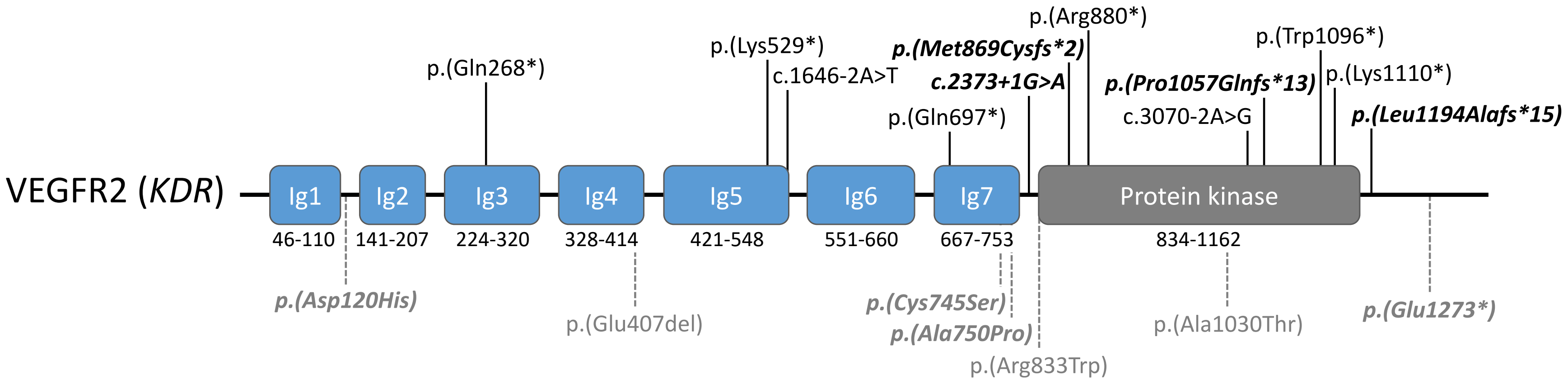


**Figure S1. Variants identified in vascular endothelial growth factor receptor 2 (VEGFR2; *KDR*: NM_002253.2), in patients with TOF or other conotruncal defects.**

Loss-of-function variants (black, top); in-frame deletion, missense variants, and stopgain variant in penultimate exon (gray, bottom).

Variants from this study (Tables S3 and S5) are in ***bold/italics***.

Variants reported in Jin et al.[28], Table S7: p.(Lys529*), c.1646-2A>T.

Variants reported in Reuter et al.[23], Figure 1: p.(Arg880*), p.(Trp1096*), p.(Glu407del), p.(Arg833Trp), p.(Ala1030Thr).

Variants reported in Morton et al.[24], Table S4 (some probands may also be reported in / overlap those reported by Jin et al.): p.(Gln268*), p.(Lys529*), c.1646-2A>T, p.(Gln697*), c.3070-2A>G, p.(Lys1110*).

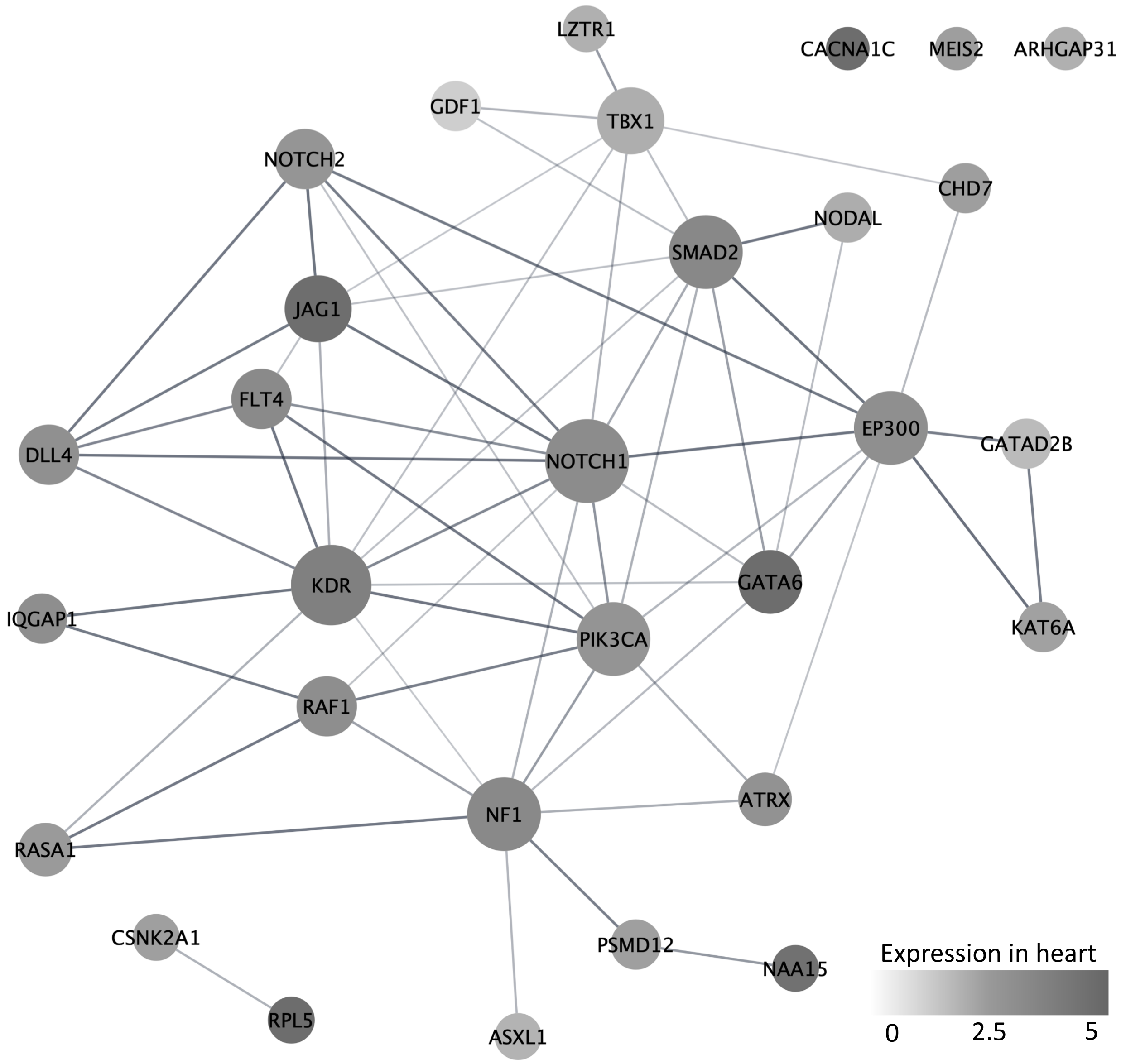


**Figure S2. Confirmed and candidate genes, identified for tetralogy of Fallot (two cohorts), encode functionally interacting proteins – extended network.** In addition to using rare high-impact variant data from the n = 811 TOF exomes from EGAS00001003302[1] (Tables S2, S3, Figure 2), this network also considers likely pathogenic loss-of-function variants available from an independent sample of n = 424 TOF exomes from Jin et al. [28] (Table S4). Network analysis was performed using Cytoscape, STRING and the total 30 CHD genes with clinically relevant variants identified in the two samples (Tables S2, S3, S4; STRING interaction enrichment *p* value = 1.0E-15). Node sizes (circles) represent the connectivity (numbers of edges to other proteins). Node greyscales represent the degree of expression from the Gene Expression Omnibus (GEO) database (heart tissue). Edge widths represent the confidence (strength of data support). VEGFR2 (gene *KDR*) and NOTCH1 form central nodes within the network, each connecting directly with 11 or 12 other proteins, respectively. NOTCH2 adds 5 interactions to the extended network, compared to the one presented in Figure 2, for the original n = 811 probands with TOF.

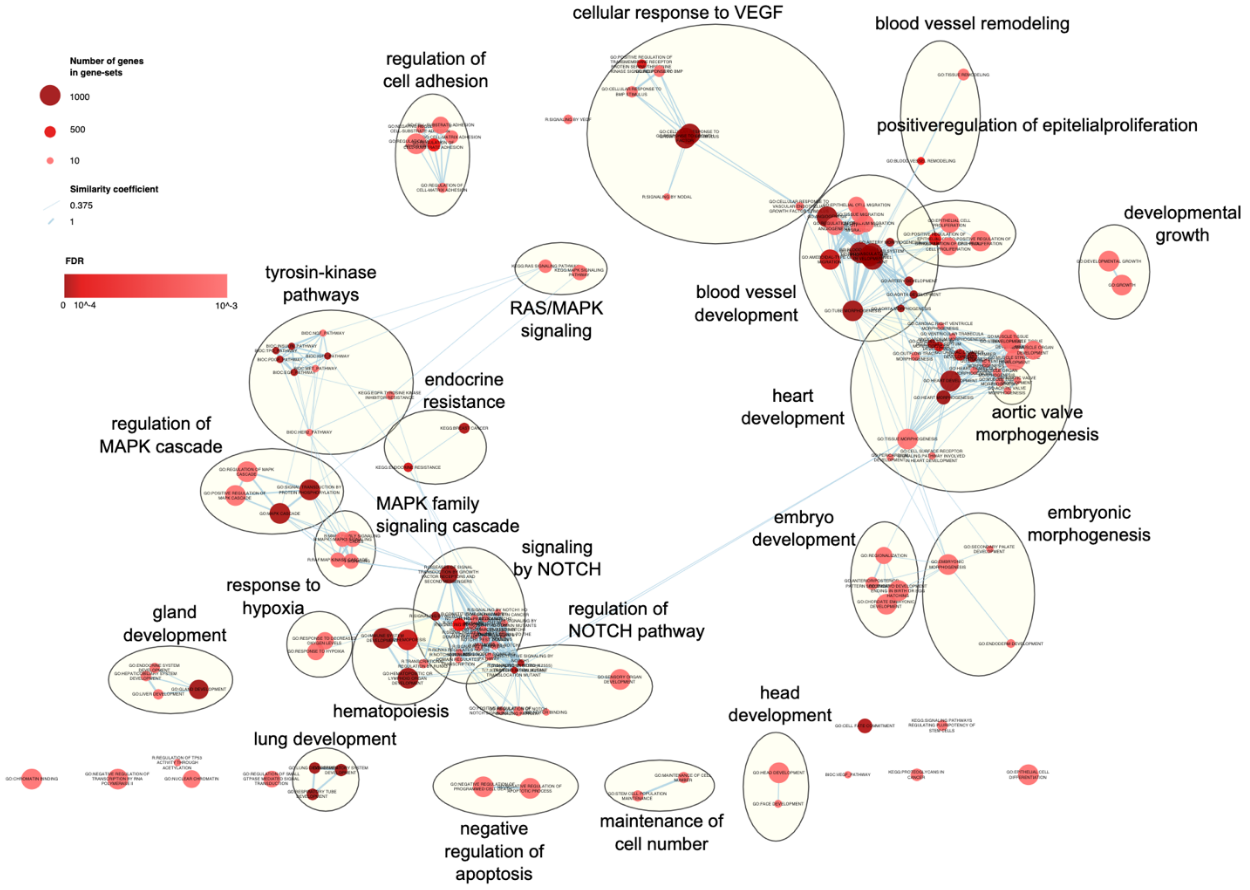


**Figure S3. Pathway enrichment map of confirmed genes (n = 23) and emerging candidate genes (n = 3) for CHD/TOF identified in n = 811 individuals with TOF**. P values and odd ratios are reported in Table S8 (provided as a separate file).
